# Association Between Neck Pain and Osteoarthritis: A Bidirectional Mendelian Randomisation Study

**DOI:** 10.1101/2025.10.13.25337861

**Authors:** Ivan Shirinsky, Valery Shirinsky, Elisaveta Alexandrovitch, Denis Plotnikov

**Affiliations:** Laboratory for the Study of Multimorbidity in Rheumatic Diseases, Research Institute of Internal and Preventive Medicine - Branch of the Institute of Cytology and Genetics, Siberian Branch of Russian Academy of Sciences, 175/1 B. Bogatkova Street, Novosibirsk 630089, Russia; Federal State Autonomous Educational Institution of Higher Education “Novosibirsk National Research State University”, Ministry of Science and Higher Education of the Russian Federation, 630090, Novosibirsk, Russian Federation; Central Research Laboratory, Kazan State Medical. University, 49 Butlerov str., 420012 Kazan, Russia

## Abstract

**Background:** Neck pain is one of the most prevalent musculoskeletal complaints and has recently been recognized as a frequent comorbidity of osteoarthritis (OA). Whether this association reflects shared biological mechanisms or bidirectional influences remains unclear.

**Objectives:** To investigate the bidirectional associations between genetic liability to neck pain and OA at three major joint sites (knee, hand, and hip) using Mendelian randomization (MR).

**Methods:** A two-sample bidirectional MR study was conducted using summary-level data from large genome-wide association studies of neck pain (579,152 participants from the Million Veteran Program) and of knee, hand, and hip OA (up to 1.3 million participants from the international Osteoarthritis Consortium). The primary analysis was random-effects inverse-variance weighted (IVW). The additional analyses comprised of multiple pleiotropy-robust sensitivity methods (weighted median, mode-based estimators, MR-Egger, MR-PRESSO, MR-Lasso).

**Results:** Genetic liability to neck pain was positively associated with knee OA (IVW OR = 1.24, 95 % CI 1.10-1.41, p < 0.001) and showed a weaker, non-significant association with hand OA after correction for multiple testing (OR = 1.41, 95 % CI 1.03-1.95, p = 0.031). No association was observed with hip OA. In the reverse direction, genetic liability to knee (OR = 1.07, 95 % CI 1.03-1.11, p < 0.001) and hip OA (OR = 1.05, 95 % CI 1.02-1.07, p < 0.001) were associated with increased risk of neck pain, whereas hand OA showed no effect. Sensitivity analyses yielded consistent results with no evidence of directional pleiotropy.

**Conclusions:** Genetic predisposition to neck pain was associated with a higher risk of knee OA, while genetic predisposition to knee and hip OA was associated with higher risk of neck pain. These bidirectional associations suggest overlapping biological pathways that warrant confirmation in independent studies.

## Background

Osteoarthritis (OA) is one of the leading causes of disability worldwide (1). The pathogenesis of pain, which is the cardinal symptom of OA, remains incompletely understood. Growing evidence suggests that comorbidities can act as important predictors of pain severity and disease burden in OA (2). Therefore, investigating the impact of specific comorbidities on OA, as well as clarifying the nature of their associations with OA, may provide novel insights into disease mechanisms and open new avenues for prevention and management.

In recent years, neck pain has emerged as a frequent comorbidity in individuals with OA. Chen et al. demonstrated an association between neck pain and OA (3). Given the high prevalence of neck pain in the general population (4), this association is of particular importance. Moreover, neck pain often develops at younger ages (5), which may pave the way for later OA, making it a potentially attractive target for early preventive strategies. A biologically plausible mechanism linking neck pain with subsequent OA symptoms is central sensitization, whereby persistent nociceptive input from the cervical spine enhances central nervous system responsiveness and lowers the threshold for musculoskeletal pain. Such sensitization has also been implicated in OA, where pain severity frequently exceeds the degree of structural joint damage (6).

Conversely, OA may predispose individuals to chronic pain in distant locations through alterations in gray matter (7).

While evidence for an association between neck pain and OA comes from an observational study (3), an alternative approach to address such questions is Mendelian randomization (MR), an analytical method that uses genetic variants as instrumental variables to investigate potential causal relationships. By leveraging the random allocation of genetic variants at conception, MR studies can minimize confounding and reverse causation that often limit observational research (8).

We therefore conducted a bidirectional MR study to test the hypotheses that genetically predicted neck pain increases the risk of knee, hip, and hand OA, and conversely, that genetically predicted OA at these sites increases the risk of neck pain.

## Methods

### Study design

We carried out a two-sample, bidirectional MR study using large-scale GWAS summary data for neck pain and for knee, hip, and hand OA (see Datasets for details). MR relies on three core assumptions:

1. Relevance - the genetic variants used as instruments are robustly associated with the exposure. 2. Independence - the instruments are not associated with confounders of the exposure-outcome relationship. 3. Exclusion restriction - the instruments affect the outcome only through the exposure, not through alternative pathways (8). Analyses were performed in both directions: Neck pain → OA, testing whether genetic liability to neck pain increases the risk of OA at specific joint sites. OA → neck pain, testing whether genetic liability to OA influences the risk of neck pain.

This bidirectional design enables to assess not only the presence but also the directionality of any potential relationships.

### Datasets

The characteristics of the datasets used for the analyses are summarized in Supplementary Table S1.

Neck pain. GWAS summary statistics for neck pain were obtained from the U.S. Department of Veterans Affairs (VA) Million Veteran Program, one of the largest biobank-based resources. Eligible participants were active users of the Veterans Health Administration who could provide informed consent; recruitment leveraged VA administrative lists for mailed invitations and clinic-based enrollment across multiple VA facilities, with written consent/HIPAA authorization, a baseline survey, and a blood draw, and linkage to longitudinal electronic health records and national VA databases. MVP was designed as a nationwide resource targeting ∼1 million enrollees (9,10). Neck pain was defined from VA EHRs using the phecode for cervicalgia, where a phecode is a curated grouping of ICD-9/ICD-10 diagnosis codes that aggregates clinically related codes into a reproducible phenotype (11). Cases required ≥2 phecode-mapped encounters and controls had none; analyses were restricted to ancestry groups with ≥200 cases per population (10).The analysis included 579,152 participants (123,311 cases and 455,841 controls; 21.3% cases), representing a multi-ancestry population (European, African American/Afro-Caribbean, Hispanic/Latino, and East Asian). Genotyping was performed using genome-wide arrays, and imputation yielded ∼44.3 million SNPs. Analyses were adjusted for age, sex, and 10 population-specific genetic principal components to account for potential confounding by ancestry. Full summary statistics are available through the GWAS Catalog (https://www.ebi.ac.uk/gwas, accession GCST90480571).

Osteoarthritis (OA). For the summary statistics for knee, hand, and hip OA we used data from the international Osteoarthritis Consortium (12). These large-scale meta-analyses combined data across multiple countries (Australia, China, Denmark, Estonia, Finland, Germany, Greece, Iceland, Japan, Netherlands, Norway, Sweden, United Kingdom, and United States), with sample sizes ranging from 890,465 (hand OA) to 1,316,500 (knee OA). The number of cases and controls was 172,256/1,144,244 for knee OA, 40,904/849,561 for hand OA, and 97,328/1,055,379 for hip OA. Case proportions ranged from 4.6% (hand OA) to 13.1% (knee OA). Ancestry composition was multi-ethnic, but the majority of participants were of European descent (85-96%). Imputation yielded between ∼19.6 and 24.3 million SNPs across phenotypes. Association testing was performed using logistic regression under an additive genetic model, with adjustment for age, sex, and principal components (or proxy measures, such as county of birth in Iceland), and results were corrected for residual stratification using LD score regression. Full summary statistics were retrieved from the GWAS Catalog (https://www.ebi.ac.uk/gwas; accessions GCST90566800, GCST90566797, and GCST90566798 for knee, hand, and hip OA, respectively).

We did not include all OA (a mixed phenotype of osteoarthritis at different joint sites) in our analyses, as knee, hip, and hand OA represent distinct disease phenotypes with potentially different underlying mechanisms (13). We also excluded spinal OA, as it is challenging to examine as an independent phenotype; it frequently co-occurs with disc degeneration, facet joint disease, and neuropathic changes, which share overlapping clinical and imaging characteristics. Moreover, there is no widely accepted research definition of spinal OA, as highlighted by a recent international Delphi consensus study (14).

### Genetic instument selection

We selected genetic variants that reached genome-wide significance (p < 5 × 10⁻⁸) in the exposure GWASs. To retain only independent signals, we applied linkage disequilibrium (LD) clumping at a threshold of r² < 0.001 within a 10 Mb window, using the European ancestry reference panel from the 1000 Genomes Project as the reference. The resulting set of independent genome-wide significant SNPs was carried forward for use as genetic instruments in Mendelian randomization analyses. The strength of the selected genetic instruments was evaluated using F-statistics. If values of F-statistics were below 10, the instrument was considered weak.

### Statistical analyses methods

The primary method of estimating the effects of each exposure on outcome was random-effects inverse variance weighted (IVW)(15). This method implies calculation of ratio of SNP-outcome association and SNP-exposure association (Wald raio) and pooling the results for each exposure-outcome pair. The random effects are used to control for between SNP heterogeneity. Because both the exposure and outcome were binary traits and effect estimates were expressed as log odds ratios (logOR), the overall estimate was scaled by multiplying with 0.693 (i.e., log(2)) to represent the change in outcome per two-fold increase in the binary exposure, as suggested by Burgess and Labrecque (16).

### Sensitivity analyses

As the standard IVW method requires strong assumptions (all genetic variants are valid instruments with no horizontal pleiotropy), we performed several additional analyses that are less sensitive to violations of these assumptions.

The weighted median method was used to provide consistent association estimates even if up to 50% of the weight comes from invalid instruments (17). The simple mode and weighted mode estimators were applied to identify association estimates under the assumption that the largest cluster of instruments represents valid variants (18). MR-Egger regression was used for two complementary purposes: (i) to obtain a pleiotropy-robust MR association estimate (the slope), which remains consistent under the InSIDE assumption even if instruments have directional pleiotropy, and (ii) to test for unbalanced horizontal pleiotropy via the intercept, where a non-zero intercept (p < 0.05) indicates directional pleiotropy (19). We further applied MR-PRESSO to detect and correct for outlier variants that may bias MR association estimates (20), and MR-Lasso as a robust regression framework that down-weights the influence of pleiotropic SNPs (21).

To assess the influence of individual instruments, we performed leave-one-out analyses, excluding each SNP in turn and recalculating the association estimate to identify whether results were driven by any single variant (22). Heterogeneity among instrument-specific estimates was assessed using the Cochran’s Q statistic under the IVW framework, while horizontal pleiotropy was evaluated using the intercept from MR-Egger regression. As we performed six primary comparisons (neck pain with knee, hip, and hand OA in both directions), we applied a Bonferroni correction to account for multiple testing, setting the threshold for statistical significance at p < 0.05/6 = 0.0083. This research was conducted using publicly available summary-level GWAS data. Ethical approval and participant consent had been obtained in the original studiens contributing to these datasets. As no individual-level data were used, no additional ethical approval was required for the present analyses. The study was reported in accordance with the STROBE-MR guidelines for strengthening the reporting of Mendelian randomization studies (23). All analyses were performed in R (version 4.4.1; (24)).

We used the following packages for Mendelian randomization analyses: TwoSampleMR (25) for instrument selection, harmonization, and core MR analyses; MR-PRESSO (26) to detect and correct for horizontal pleiotropy; MendelianRandomization (27) for MR-Lasso estimation.

Additional data wrangling and visualization were conducted with the *tidyverse* packages (28). For reporting and table formatting, we used *flextable* (29) and *officedown* (30).

## Results

### Selection of genetic instruments

The characteristics of the genetic instruments are summarized in Supplementary Tables S2-S3. After genome-wide significance filtering and LD clumping, we retained 7 SNPs for neck pain, 128 SNPs for knee OA, 18 SNPs for hand OA, and 128 SNPs for hip OA. All selected instruments had mean F-statistics greater than 10, with values ranging from 29.9 to 199.5.

### Heterogeneity and pleiotropy

Table 1 summarizes the heterogeneity statistics (Cochran’s Q test) and MR-Egger intercepts across all exposure-outcome pairs. Evidence of heterogeneity was observed for the associations of neck pain with knee, hand, and hip OA, as well as for the associations of knee, hand, and hip OA with neck pain (all Q-test p < 0.05). In contrast, MR-Egger intercepts were not significantly different from zero for any exposure-outcome pair, indicating no evidence of unbalanced horizontal pleiotropy.

**Table 1.** Heterogeneity statistics and MR Egger intercepts across exposure-outcome pairs.

| Exposure | Outcome | Cochran's Q test |  | Egger test |  |
| --- | --- | --- | --- | --- | --- |
|  |  | Q value | P | Egger intercept | P |
| Neck pain | Knee OA | 16.43 | 0.006 | 0.002 | 0.874 |
| Neck pain | Hand OA | 11.61 | 0.040 | 0.010 | 0.698 |
| Neck pain | Hip OA | 18.16 | 0.003 | 0.011 | 0.504 |
| Knee OA | Neck pain | 303.11 | 0.000 | 0.004 | 0.199 |
| Hand OA | Neck pain | 38.19 | 0.001 | 0.007 | 0.596 |
| Hip OA | Neck pain | 262.05 | 0.000 | 0.002 | 0.520 |

Figure 1 presents the associations of genetically predicted neck pain with genetically predicted knee, hand, and hip OA assessed using multiple Mendelian randomization methods. Supporting diagnostic plots, including scatter, per-SNP forest, and funnel plots, are provided in Supplementary Figures S1, S3, and S5.

**Figure 1.**
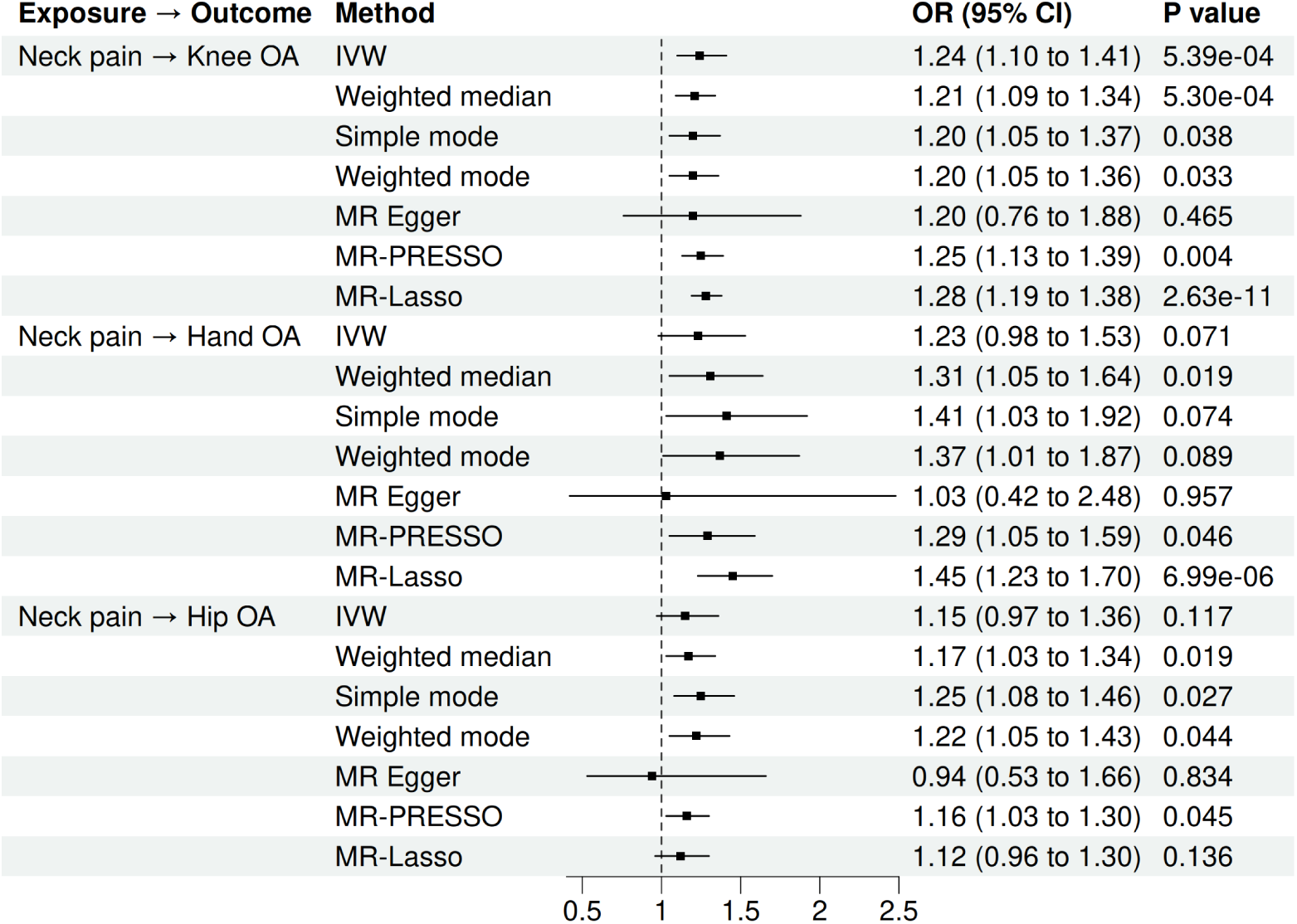
Associations of genetically predicted neck pain with osteoarthritis.

For knee OA, the IVW analysis indicated a significant positive association (OR = 1.24, 95% CI: 1.10-1.41, p < 0.001), which remained significant after Bonferroni correction. Sensitivity analyses, including the weighted median, mode-based estimators, MR-PRESSO, and MR-Lasso, produced consistent results. MR-Egger regression suggested a positive direction of effect, though it did not reach statistical significance.

For hand OA, the IVW method also demonstrated a positive association (OR = 1.41, 95% CI: 1.03- 1.95, p = 0.031). Although supported by the weighted median and MR-Lasso approaches, this association did not reach the Bonferroni-adjusted significance threshold. Mode-based methods indicated effects of similar magnitude but with wider uncertainty intervals. MR-Egger regression again yielded a non-significant estimate.

For hip OA, the IVW analysis did not identify a significant association (OR = 1.05, 95% CI: 0.97- 1.14, p = 0.177). Sensitivity analyses using the weighted median, mode-based methods, MR-PRESSO, and MR-Lasso suggested effects in the same direction but without statistical significance. MR-Egger regression also indicated no effect.

Leave-one-out analyses of neck pain on OA did not identify any individual variants that disproportionately influenced the overall association estimates (Supplementary figure S7).

### Associations of genetically predicted OA with genetically predicted neck pain

Figure 2 presents the associations of genetically predicted knee, hand, and hip OA with genetically predicted neck pain evaluated using multiple Mendelian randomization methods. Supplementary Figures S2-S4 present the corresponding scatter plots and per-SNP forest plots for these analyses, while Supplementary Figure S6 shows funnel plots assessing asymmetry.

**Figure 2.**
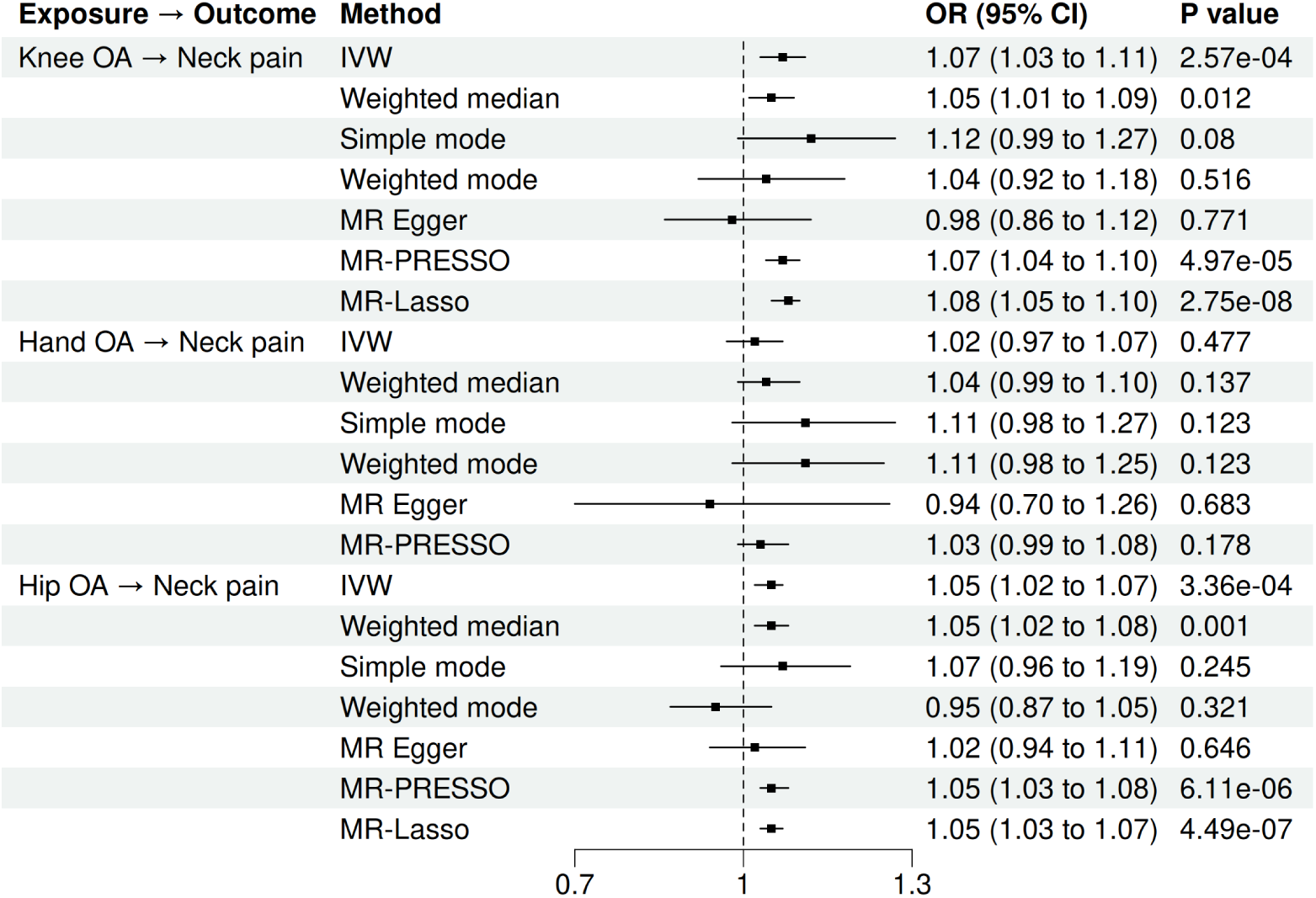
Associations of genetically predicted osteoarthritis with neck pain.

For knee OA, the IVW analysis indicated a significant positive association (OR = 1.07, 95% CI: 1.03-1.11, p < 0.001), which remained significant after Bonferroni correction. The weighted median method gave a consistent effect estimate, while mode-based estimators were less precise. Robust methods, including MR-PRESSO and MR-Lasso, strongly supported this association. MR-Egger regression suggested no effect.

For hand OA, the IVW analysis did not identify a significant association with neck pain (OR = 1.02, 95% CI: 0.97-1.07, p = 0.477). Sensitivity analyses, including the weighted median, mode-based methods, MR-PRESSO, and MR-Lasso, produced similar null results. MR-Egger regression also indicated no effect.

For hip OA, the IVW analysis showed a significant positive association (OR = 1.05, 95% CI: 1.02- 1.07, p < 0.001), which remained significant after Bonferroni correction. This result was supported by the weighted median, MR-PRESSO, and MR-Lasso approaches, whereas mode-based estimators yielded less consistent findings. MR-Egger regression again suggested no effect.

Leave-one-out analyses (Supplementary Figure S8) supported the small but significant effects for hip and knee OA and the null for hand OA.

## Discussion

In this Mendelian randomization study, we investigated the bidirectional associations between genetically predicted neck pain and osteoarthritis at three sites (knee, hand, and hip). We found consistent evidence that genetically predicted neck pain was associated with an increased risk of knee OA, with results robust across multiple sensitivity analyses and after Bonferroni correction. A suggestive but weaker association was observed with hand OA, which did not remain significant after correction for multiple testing, while no association was found with hip OA. In the reverse direction, genetically predicted knee and hip OA were also associated with a higher risk of neck pain, although the effect sizes were smaller than those observed for neck pain on OA. No evidence of association was found between hand OA and neck pain.

This study has several limitations. First, the number of genome-wide significant variants available for neck pain was modest, which may limit statistical power to detect small effects. Second, although we applied multiple pleiotropy-robust sensitivity methods, residual horizontal pleiotropy cannot be completely excluded. Third, both neck pain and OA phenotypes were derived from large-scale GWAS meta-analyses that rely partly on self-reported or electronic health record data, which may introduce misclassification and heterogeneity across studies. Finally, Mendelian randomization estimates reflect the effects of lifelong genetic liability rather than the clinical or modifiable aspects of neck pain and OA, and thus should not be directly equated with outcomes of clinical interventions.

These Mendelian randomization analyses suggest directional associations of genetic liability to neck pain with knee and hand osteoarthritis, and of knee and hip osteoarthritis with neck pain, with varying strength of evidence across directions. This pattern may indicate partially shared biological and pain-related mechanisms rather than direct causal effects.

A biologically plausible pathway linking neck pain and osteoarthritis may be central sensitization, whereby persistent nociceptive input from the cervical spine lowers the threshold for musculoskeletal pain (6). Conversely, the reverse association—where osteoarthritis predisposes to neck pain—may reflect secondary central sensitization caused by persistent peripheral nociceptive input from joints affected by OA. Furthermore, chronic neck pain is characterized by impaired biomechanics, including altered proprioception and disturbed gait patterns, which may alter joint loading and movement efficiency (31). Neck pain has also been associated with reduced gait speed (32) while slower gait has been linked to higher osteoarthritis risk (33). Taken together, these neurophysiological and biomechanical mechanisms may explain the bidirectional associations observed in this study.

Common pleiotropic pathways could predispose individuals to both conditions independently. Our findings, together with overlapping biological plausibility, may indicate partial genetic correlation rather than mutual causation. Future studies employing multivariable Mendelian randomization could help clarify whether the associations still observeed after adjusting for shared risk factors, such as body mass index, inflammatory markers, or general pain sensitivity. Additionally, network MR or phenome-wide causal modeling (34) may further separate shared etiological pathways from potential feedback effects. The present findings suggest the clinical importance of recognizing neck pain as part of the osteoarthritis spectrum. Identifying and addressing neck pain, particularly in younger adults, may help clinicians detect individuals at risk for multisite pain and functional decline earlier in the disease course and initiate preventive strategies aimed at neck pain, if this association is further supported by independent studies.

The generalisability of these findings is limited by the predominantly European ancestry of the GWAS samples and by potential differences between genetically proxied traits and their clinically observed phenotypes.

In conclusion, this bidirectional Mendelian randomization study provides evidence that genetic liability to neck pain is associated with an increased risk of knee OA, while genetic liability to knee and hip OA is associated with a higher risk of neck pain. Further research integrating genetic, neurophysiological, and biomechanical data across diverse populations is warranted to clarify these relationships and to inform potential strategies for early risk identification and prevention.

## Data Availability

No individual-level data were accessed or used in this analysis.
All data used in this study were obtained from publicly available GWAS summary statistics.GWAS summary statistics for neck pain were obtained from the U.S. Department of Veterans Affairs Million Veteran Program and are accessible through the GWAS Catalog under accession GCST90480571 (https://www.ebi.ac.uk/gwas/studies/GCST90480571). GWAS summary statistics for knee, hand, and hip osteoarthritis (OA) were obtained from the international Osteoarthritis Consortium and are available through the GWAS Catalog under accessions GCST90566800 (https://www.ebi.ac.uk/gwas/studies/GCST90566800), GCST90566797 (https://www.ebi.ac.uk/gwas/studies/GCST90566797), and GCST90566798 (https://www.ebi.ac.uk/gwas/studies/GCST90566798), respectively.
The custom scripts contain institution-specific file paths and are available from the corresponding author upon reasonable request.

https://www.ebi.ac.uk/gwas/studies/GCST90566800

https://www.ebi.ac.uk/gwas/studies/GCST90566797

https://www.ebi.ac.uk/gwas/studies/GCST90566798

## Acknowledgements

None

## Data and data sharing

No individual-level data were accessed or used in this analysis. All data used in this study were obtained from publicly available GWAS summary statistics.GWAS summary statistics for neck pain were obtained from the U.S. Department of Veterans Affairs Million Veteran Program and are accessible through the GWAS Catalog under accession GCST90480571 (https://www.ebi.ac.uk/gwas/studies/GCST90480571). GWAS summary statistics for knee, hand, and hip osteoarthritis (OA) were obtained from the international Osteoarthritis Consortium and are available through the GWAS Catalog under accessions GCST90566800 (https://www.ebi.ac.uk/gwas/studies/GCST90566800), GCST90566797 (https://www.ebi.ac.uk/gwas/studies/GCST90566797), and GCST90566798 (https://www.ebi.ac.uk/gwas/studies/GCST90566798), respectively. The analyses were conducted using standard R packages. The custom scripts contain institution-specific file paths and are available from the corresponding author upon reasonable request.

---

**Supplementary Table S1.** GWAS sample composition and study characteristics.

| Variable | Neck pain<br>(GCST90480571) | Knee OA (GCST90566800) | Hand OA (GCST90566797) | Hip OA (GCST90566798) |
| --- | --- | --- | --- | --- |
| N_total | 579,152 | 1,316,500 | 890,465 | 1,152,707 |
| N_cases /<br>N_controls (%<br>cases) | 123,311 / 455,841<br>(21.3%) | 172,256 / 1,144,244 (13.1%) | 40,904 / 849,561 (4.6%) | 97,328 / 1,055,379 (8.4%) |
| African ancestry | 108,180 (18.7%) | 24,911 (1.9%) | 15,078 (1.7%) | 22,751 (2.0%) |
| Hispanic / Latin<br>ancestry | 53,954 (9.3%) | 3,426 (0.3%) | 3,713 (0.4%) | 3,774 (0.3%) |
| East Asian<br>ancestry | 6,280 (1.1%) | 132,202 (10.0%) | 484 (0.1%) | 2,684 (0.2%) |
| European ancestry | 410,738 (70.9%) | 1,127,213 (85.6%) | 853,901 (95.9%) | 1,107,086 (96.0%) |
| South Asian<br>ancestry | 0 (0.0%) | 19,864 (1.5%) | 8,839 (1.0%) | 7,997 (0.7%) |
| Other ancestry | 0 (0.0%) | 8,884 (0.7%) | 8,450 (0.9%) | 8,415 (0.7%) |
| Ethnicity (overall) | Multi-ancestry: Eur., Afr.-<br>Am., Hisp., East Asian | Multi-ancestry: Eur., East Asian, Afr./Af-Am.,<br>South Asian, Hisp., Admixed | Multi-ancestry: Eur., East Asian, Afr./Af-Am., South Asian, Hisp., Admixed | Multi-ancestry: Eur., East Asian, Afr./Af-Am.,<br>South Asian, Hisp., Admixed |
| Countries (ISO-2) | US | AU, CN, DK, EE, FI, DE, GR, IS, JP, NL, NO,<br>SE, UK, US | AU, CN, DK, EE, FI, DE, GR, IS, JP, NL,<br>NO, SE, UK, US | AU, CN, DK, EE, FI, DE, GR, IS, JP, NL, NO,<br>SE, UK, US |
| GWAS type | Primary GWAS (multi-ancestry) | Meta-analysis (multi-ancestry) | Meta-analysis (multi-ancestry) | Meta-analysis (multi-ancestry) |
| Phenotype definition | VA EHR cervicalgia phecode (curated ICD-9/10 grouping) | Osteoarthritis of the knee (ascertained using a combination of self-report, hospital diagnosis, and radiographic criteria) | Osteoarthritis of the hand (ascertained using a combination of self-report, hospital diagnosis, and radiographic criteria) | Osteoarthritis of the hip (ascertained using a combination of self-report, hospital diagnosis, and radiographic criteria) |
| Unit | logOR | logOR | logOR | logOR |
| Platform | Genome-wide array | Affymetrix, Illumina | Affymetrix, Illumina | Affymetrix, Illumina |
| Imputed SNPs | 44,300,000 | 24,344,537 | 19,591,543 | 20,097,305 |
| Access site (NHGRI-EBI) | GCST90480571.tsv.gz | GCST90566800.tsv.gz | GCST90566797.tsv.gz | GCST90566798.tsv.gz |
| Access date | 2025-08-13 | 2025-08-13 | 2025-08-13 | 2025-08-13 |
| Reference | Science 2024; PMID: 39024449 (Verma et al.) | Nature 2025; PMID: 40205036 (Hatzikotoulas et al.) | Nature 2025; PMID: 40205036 (Hatzikotoulas et al.) | Nature 2025; PMID: 40205036 (Hatzikotoulas et al.) |

**Supplementary Table S2.** Per-SNP F-statistics (exposure instruments).

| exposure | SNP | beta.exposure | se.exposure | F_statistic | p |
| --- | --- | --- | --- | --- | --- |
| neck pain | rs1476535 | 0.03718 | 0.005422 | 47.033 | 6.98e-12 |
| neck pain | rs11663694 | 0.03718 | 0.005569 | 44.573 | 2.45e-11 |
| neck pain | rs10891490 | -0.03247 | 0.005406 | 36.063 | 1.91e-09 |
| neck pain | rs8037430 | 0.03273 | 0.005839 | 31.418 | 2.08e-08 |
| neck pain | rs2684618 | -0.08526 | 0.015320 | 30.985 | 2.60e-08 |
| neck pain | rs564957276 | -0.10530 | 0.019120 | 30.316 | 3.67e-08 |
| neck pain | rs769670 | -0.03053 | 0.005579 | 29.943 | 4.45e-08 |
| knee OA | rs9940278 | 0.05360 | 0.004183 | 164.180 | 1.38e-37 |
| knee OA | rs11731421 | 0.05190 | 0.004383 | 140.200 | 2.41e-32 |
| knee OA | rs4606747 | -0.05120 | 0.004482 | 130.480 | 3.22e-30 |
| knee OA | rs12310519 | 0.06020 | 0.005742 | 109.920 | 1.02e-25 |
| knee OA | rs429358 | -0.06220 | 0.006478 | 92.191 | 7.87e-22 |
| knee OA | rs3843750 | -0.04440 | 0.004792 | 85.834 | 1.96e-20 |
| knee OA | rs10048129 | 0.04640 | 0.005113 | 82.338 | 1.15e-19 |
| knee OA | rs876122 | 0.06080 | 0.006857 | 78.610 | 7.57e-19 |
| knee OA | rs10859587 | 0.03810 | 0.004309 | 78.166 | 9.47e-19 |
| knee OA | rs148024643 | 0.04940 | 0.005633 | 76.909 | 1.79e-18 |
| knee OA | rs35697691 | 0.07820 | 0.008979 | 75.846 | 3.07e-18 |
| knee OA | rs1078301 | 0.04190 | 0.004866 | 74.139 | 7.28e-18 |
| knee OA | rs6510645 | 0.03600 | 0.004279 | 70.766 | 4.02e-17 |
| knee OA | rs2820436 | -0.03860 | 0.004623 | 69.728 | 6.81e-17 |
| knee OA | rs12911554 | -0.03580 | 0.004357 | 67.526 | 2.08e-16 |
| knee OA | rs2867109 | -0.05070 | 0.006215 | 66.550 | 3.41e-16 |
| knee OA | rs9533090 | -0.03440 | 0.004224 | 66.321 | 3.83e-16 |
| knee OA | rs4425336 | 0.03830 | 0.004872 | 61.810 | 3.78e-15 |
| knee OA | rs12214906 | -0.03240 | 0.004198 | 59.569 | 1.18e-14 |
| knee OA | rs10878346 | -0.03870 | 0.005026 | 59.285 | 1.36e-14 |
| knee OA | rs11749736 | 0.03600 | 0.004832 | 55.518 | 9.26e-14 |
| knee OA | rs3774364 | 0.03170 | 0.004257 | 55.450 | 9.59e-14 |
| knee OA | rs1059698 | -0.03530 | 0.004749 | 55.241 | 1.07e-13 |
| knee OA | rs10974438 | -0.03170 | 0.004349 | 53.131 | 3.12e-13 |
| knee OA | rs1325032 | 0.03190 | 0.004410 | 52.335 | 4.68e-13 |
| knee OA | rs2076545 | 0.03050 | 0.004221 | 52.200 | 5.01e-13 |
| knee OA | rs450466 | -0.03050 | 0.004233 | 51.911 | 5.81e-13 |
| knee OA | rs57525841 | 0.03620 | 0.005094 | 50.491 | 1.20e-12 |
| knee OA | rs8015712 | 0.03350 | 0.004718 | 50.406 | 1.25e-12 |
| knee OA | rs2015110 | 0.03330 | 0.004691 | 50.386 | 1.26e-12 |
| knee OA | rs1426371 | -0.03380 | 0.004777 | 50.067 | 1.49e-12 |
| knee OA | rs11168351 | -0.03090 | 0.004370 | 50.008 | 1.53e-12 |
| knee OA | rs10995385 | 0.03340 | 0.004759 | 49.256 | 2.25e-12 |
| knee OA | rs13107325 | 0.06690 | 0.009612 | 48.446 | 3.40e-12 |
| knee OA | rs2242279 | 0.03120 | 0.004501 | 48.055 | 4.14e-12 |
| knee OA | rs12925886 | 0.03340 | 0.004857 | 47.294 | 6.11e-12 |
| knee OA | rs359974 | 0.04030 | 0.005862 | 47.265 | 6.20e-12 |
| knee OA | rs887258 | -0.03680 | 0.005379 | 46.802 | 7.85e-12 |
| knee OA | rs35021943 | -0.03410 | 0.005046 | 45.673 | 1.40e-11 |
| knee OA | rs1431402 | -0.03830 | 0.005697 | 45.203 | 1.78e-11 |
| knee OA | rs1515405 | -0.02890 | 0.004299 | 45.186 | 1.79e-11 |
| knee OA | rs2738293 | -0.03230 | 0.004818 | 44.944 | 2.03e-11 |
| knee OA | rs3118919 | -0.03490 | 0.005214 | 44.798 | 2.18e-11 |
| knee OA | rs79057214 | -0.04590 | 0.006898 | 44.281 | 2.84e-11 |
| knee OA | rs2443701 | -0.03530 | 0.005311 | 44.181 | 2.99e-11 |
| knee OA | rs2521348 | 0.02800 | 0.004230 | 43.810 | 3.62e-11 |
| knee OA | rs55658362 | -0.07010 | 0.010660 | 43.251 | 4.82e-11 |
| knee OA | rs12155373 | -0.02930 | 0.004460 | 43.166 | 5.03e-11 |
| knee OA | rs11042731 | 0.03220 | 0.004912 | 42.979 | 5.53e-11 |
| knee OA | rs963292 | -0.02770 | 0.004230 | 42.881 | 5.82e-11 |
| knee OA | rs9397078 | -0.04000 | 0.006145 | 42.374 | 7.54e-11 |
| knee OA | rs11614330 | -0.02830 | 0.004355 | 42.232 | 8.10e-11 |
| knee OA | rs10831476 | -0.03490 | 0.005423 | 41.422 | 1.23e-10 |
| knee OA | rs2568956 | 0.03250 | 0.005064 | 41.186 | 1.38e-10 |
| knee OA | rs148449802 | -0.03630 | 0.005670 | 40.980 | 1.54e-10 |
| knee OA | rs935166 | -0.02880 | 0.004516 | 40.665 | 1.81e-10 |
| knee OA | rs77165542 | -0.08440 | 0.013260 | 40.538 | 1.93e-10 |
| knee OA | rs11001784 | 0.02790 | 0.004391 | 40.372 | 2.10e-10 |
| knee OA | rs77131772 | 0.04080 | 0.006425 | 40.331 | 2.14e-10 |
| knee OA | rs73164496 | -0.03400 | 0.005394 | 39.727 | 2.92e-10 |
| knee OA | rs6124883 | -0.02750 | 0.004382 | 39.385 | 3.48e-10 |
| knee OA | rs11155510 | -0.02600 | 0.004157 | 39.111 | 4.00e-10 |
| knee OA | rs79950770 | -0.04020 | 0.006440 | 38.965 | 4.32e-10 |
| knee OA | rs10810601 | -0.02930 | 0.004711 | 38.680 | 4.99e-10 |
| knee OA | rs1447132 | -0.02750 | 0.004422 | 38.671 | 5.02e-10 |
| knee OA | rs805318 | -0.02580 | 0.004155 | 38.549 | 5.34e-10 |
| knee OA | rs1043801 | -0.05710 | 0.009220 | 38.351 | 5.91e-10 |
| knee OA | rs36714 | -0.03180 | 0.005140 | 38.280 | 6.13e-10 |
| knee OA | rs228768 | -0.02930 | 0.004756 | 37.948 | 7.26e-10 |
| knee OA | rs58109580 | -0.04030 | 0.006552 | 37.833 | 7.70e-10 |
| knee OA | rs113362184 | 0.02640 | 0.004295 | 37.787 | 7.89e-10 |
| knee OA | rs10915873 | 0.02970 | 0.004853 | 37.455 | 9.36e-10 |
| knee OA | rs2281997 | -0.02890 | 0.004722 | 37.454 | 9.36e-10 |
| knee OA | rs4432271 | 0.03720 | 0.006105 | 37.130 | 1.10e-09 |
| knee OA | rs10831907 | -0.02610 | 0.004288 | 37.051 | 1.15e-09 |
| knee OA | rs7350160 | -0.03760 | 0.006195 | 36.834 | 1.29e-09 |
| knee OA | rs2936427 | -0.02670 | 0.004412 | 36.622 | 1.43e-09 |
| knee OA | rs2628578 | -0.04710 | 0.007791 | 36.546 | 1.49e-09 |
| knee OA | rs17138562 | 0.04110 | 0.006804 | 36.493 | 1.53e-09 |
| knee OA | rs6090040 | 0.02720 | 0.004504 | 36.469 | 1.55e-09 |
| knee OA | rs2294098 | 0.05520 | 0.009152 | 36.378 | 1.62e-09 |
| knee OA | rs9921416 | -0.02530 | 0.004205 | 36.201 | 1.78e-09 |
| knee OA | rs28609134 | -0.02750 | 0.004583 | 36.009 | 1.96e-09 |
| knee OA | rs16977012 | 0.03060 | 0.005123 | 35.679 | 2.33e-09 |
| knee OA | rs9277928 | 0.03090 | 0.005176 | 35.634 | 2.38e-09 |
| knee OA | rs507261 | -0.02730 | 0.004585 | 35.446 | 2.62e-09 |
| knee OA | rs931794 | 0.03050 | 0.005143 | 35.176 | 3.01e-09 |
| knee OA | rs4877697 | -0.03110 | 0.005265 | 34.895 | 3.48e-09 |
| knee OA | rs10987373 | 0.02880 | 0.004887 | 34.726 | 3.80e-09 |
| knee OA | rs2731982 | -0.02540 | 0.004314 | 34.672 | 3.90e-09 |
| knee OA | rs7923649 | 0.03310 | 0.005668 | 34.103 | 5.23e-09 |
| knee OA | rs2529489 | -0.02790 | 0.004803 | 33.742 | 6.29e-09 |
| knee OA | rs35159887 | 0.02620 | 0.004515 | 33.671 | 6.53e-09 |
| knee OA | rs7160656 | -0.02550 | 0.004400 | 33.590 | 6.80e-09 |
| knee OA | rs7186541 | 0.03110 | 0.005367 | 33.575 | 6.86e-09 |
| knee OA | rs34533676 | 0.04780 | 0.008269 | 33.419 | 7.43e-09 |
| knee OA | rs7539021 | 0.02590 | 0.004484 | 33.365 | 7.64e-09 |
| knee OA | rs11742813 | 0.02960 | 0.005140 | 33.167 | 8.46e-09 |
| knee OA | rs34517439 | 0.03790 | 0.006606 | 32.918 | 9.61e-09 |
| knee OA | rs78469175 | 0.04770 | 0.008333 | 32.763 | 1.04e-08 |
| knee OA | rs7204132 | -0.02850 | 0.004982 | 32.722 | 1.06e-08 |
| knee OA | rs56283067 | 0.02650 | 0.004635 | 32.683 | 1.08e-08 |
| knee OA | rs1507465 | 0.02430 | 0.004253 | 32.645 | 1.11e-08 |
| knee OA | rs35489988 | 0.02590 | 0.004534 | 32.635 | 1.11e-08 |
| knee OA | rs73217710 | -0.04880 | 0.008546 | 32.605 | 1.13e-08 |
| knee OA | rs10744137 | 0.02820 | 0.004955 | 32.384 | 1.27e-08 |
| knee OA | rs1024896 | 0.02600 | 0.004573 | 32.330 | 1.30e-08 |
| knee OA | rs55965541 | 0.03910 | 0.006884 | 32.258 | 1.35e-08 |
| knee OA | rs7189660 | 0.02590 | 0.004564 | 32.208 | 1.38e-08 |
| knee OA | rs1530800 | -0.02400 | 0.004235 | 32.111 | 1.46e-08 |
| knee OA | rs11612013 | 0.04520 | 0.007980 | 32.083 | 1.48e-08 |
| knee OA | rs6012893 | -0.02710 | 0.004785 | 32.074 | 1.48e-08 |
| knee OA | rs10493211 | 0.02530 | 0.004469 | 32.048 | 1.50e-08 |
| knee OA | rs1895583 | -0.02740 | 0.004841 | 32.037 | 1.51e-08 |
| knee OA | rs12906125 | -0.03070 | 0.005431 | 31.957 | 1.58e-08 |
| knee OA | rs2422133 | 0.02390 | 0.004249 | 31.640 | 1.86e-08 |
| knee OA | rs141845046 | 0.06560 | 0.011660 | 31.632 | 1.86e-08 |
| knee OA | rs6567160 | 0.02760 | 0.004948 | 31.120 | 2.43e-08 |
| knee OA | rs147329802 | 0.02340 | 0.004212 | 30.862 | 2.77e-08 |
| knee OA | rs6444262 | -0.02580 | 0.004645 | 30.849 | 2.79e-08 |
| knee OA | rs61944841 | 0.02870 | 0.005182 | 30.672 | 3.06e-08 |
| knee OA | rs11030477 | 0.02750 | 0.004968 | 30.645 | 3.10e-08 |
| knee OA | rs10846657 | -0.02480 | 0.004505 | 30.309 | 3.68e-08 |
| knee OA | rs1800472 | 0.08200 | 0.014930 | 30.175 | 3.95e-08 |
| knee OA | rs1964521 | -0.02410 | 0.004391 | 30.130 | 4.04e-08 |
| knee OA | rs79970150 | 0.02680 | 0.004894 | 29.992 | 4.34e-08 |
| knee OA | rs1999659 | -0.02300 | 0.004201 | 29.971 | 4.39e-08 |
| knee OA | rs227723 | 0.02460 | 0.004498 | 29.910 | 4.52e-08 |
| hand OA | rs10851633 | 0.08440 | 0.008664 | 94.894 | 2.01e-22 |
| hand OA | rs10500759 | -0.07170 | 0.008543 | 70.447 | 4.73e-17 |
| hand OA | rs3821269 | -0.06180 | 0.008460 | 53.367 | 2.77e-13 |
| hand OA | rs2241058 | -0.07620 | 0.011610 | 43.067 | 5.29e-11 |
| hand OA | rs34255979 | -0.08880 | 0.013530 | 43.065 | 5.30e-11 |
| hand OA | rs4760611 | 0.07160 | 0.011280 | 40.305 | 2.17e-10 |
| hand OA | rs10062749 | 0.05910 | 0.009411 | 39.436 | 3.39e-10 |
| hand OA | rs10407692 | -0.06160 | 0.010000 | 37.949 | 7.26e-10 |
| hand OA | rs1366202 | -0.07520 | 0.012310 | 37.290 | 1.02e-09 |
| hand OA | rs7294636 | 0.05270 | 0.008672 | 36.932 | 1.22e-09 |
| hand OA | rs7748189 | 0.06600 | 0.011100 | 35.334 | 2.78e-09 |
| hand OA | rs72660316 | -0.07170 | 0.012190 | 34.587 | 4.08e-09 |
| hand OA | rs858335 | 0.05410 | 0.009243 | 34.258 | 4.83e-09 |
| hand OA | rs62293657 | -0.05000 | 0.008729 | 32.810 | 1.02e-08 |
| hand OA | rs17208656 | 0.04800 | 0.008607 | 31.103 | 2.45e-08 |
| hand OA | rs12618894 | -0.05140 | 0.009265 | 30.779 | 2.89e-08 |
| hand OA | rs73444590 | -0.06450 | 0.011710 | 30.329 | 3.65e-08 |
| hand OA | rs1381965 | -0.06140 | 0.011170 | 30.212 | 3.87e-08 |
| hip OA | rs12209223 | 0.11560 | 0.008185 | 199.480 | 2.71e-45 |
| hip OA | rs10843013 | 0.08670 | 0.006410 | 182.970 | 1.09e-41 |
| hip OA | rs1895062 | 0.06900 | 0.005345 | 166.660 | 3.98e-38 |
| hip OA | rs2716212 | 0.06320 | 0.005351 | 139.500 | 3.42e-32 |
| hip OA | rs17257909 | -0.06340 | 0.005532 | 131.360 | 2.06e-30 |
| hip OA | rs2673421 | -0.06030 | 0.005406 | 124.440 | 6.76e-29 |
| hip OA | rs1926872 | -0.06090 | 0.005489 | 123.100 | 1.33e-28 |
| hip OA | rs1851610 | -0.05780 | 0.005258 | 120.850 | 4.13e-28 |
| hip OA | rs2118540 | 0.05850 | 0.005335 | 120.240 | 5.61e-28 |
| hip OA | rs66989638 | 0.08100 | 0.007627 | 112.780 | 2.41e-26 |
| hip OA | rs11164653 | 0.05900 | 0.005617 | 110.330 | 8.32e-26 |
| hip OA | rs56283067 | 0.05820 | 0.005729 | 103.200 | 3.03e-24 |
| hip OA | rs11732213 | -0.06360 | 0.006618 | 92.345 | 7.28e-22 |
| hip OA | rs111844273 | 0.19030 | 0.020340 | 87.526 | 8.32e-21 |
| hip OA | rs12790261 | -0.09980 | 0.010750 | 86.146 | 1.67e-20 |
| hip OA | rs10448285 | -0.05130 | 0.005539 | 85.767 | 2.02e-20 |
| hip OA | rs77706860 | 0.06870 | 0.007478 | 84.390 | 4.06e-20 |
| hip OA | rs9941349 | 0.04860 | 0.005303 | 83.994 | 4.96e-20 |
| hip OA | rs80287694 | 0.07770 | 0.008497 | 83.629 | 5.97e-20 |
| hip OA | rs7020944 | -0.06050 | 0.006636 | 83.119 | 7.73e-20 |
| hip OA | rs1809889 | -0.05490 | 0.006084 | 81.437 | 1.81e-19 |
| hip OA | rs7814941 | -0.05960 | 0.006667 | 79.925 | 3.89e-19 |
| hip OA | rs12051660 | 0.06300 | 0.007269 | 75.126 | 4.42e-18 |
| hip OA | rs1755646 | -0.04710 | 0.005526 | 72.653 | 1.55e-17 |
| hip OA | rs79723785 | 0.15730 | 0.018920 | 69.090 | 9.41e-17 |
| hip OA | rs4794665 | -0.04580 | 0.005533 | 68.528 | 1.25e-16 |
| hip OA | rs5750495 | -0.05410 | 0.006749 | 64.262 | 1.09e-15 |
| hip OA | rs6117282 | -0.04280 | 0.005345 | 64.121 | 1.17e-15 |
| hip OA | rs788867 | 0.04420 | 0.005576 | 62.844 | 2.24e-15 |
| hip OA | rs429358 | -0.05750 | 0.007268 | 62.585 | 2.55e-15 |
| hip OA | rs9348967 | -0.05090 | 0.006455 | 62.187 | 3.12e-15 |
| hip OA | rs13107325 | 0.09270 | 0.011790 | 61.781 | 3.84e-15 |
| hip OA | rs79514988 | 0.11840 | 0.015150 | 61.042 | 5.59e-15 |
| hip OA | rs2738292 | -0.04640 | 0.005950 | 60.805 | 6.30e-15 |
| hip OA | rs12866004 | -0.04940 | 0.006368 | 60.189 | 8.62e-15 |
| hip OA | rs12636716 | -0.04440 | 0.005821 | 58.176 | 2.40e-14 |
| hip OA | rs7538503 | 0.04380 | 0.005743 | 58.169 | 2.40e-14 |
| hip OA | rs72827088 | 0.05100 | 0.006912 | 54.436 | 1.61e-13 |
| hip OA | rs12951993 | -0.04090 | 0.005564 | 54.043 | 1.96e-13 |
| hip OA | rs3780325 | 0.04670 | 0.006369 | 53.767 | 2.26e-13 |
| hip OA | rs1519038 | -0.05140 | 0.007050 | 53.160 | 3.08e-13 |
| hip OA | rs9362113 | -0.04820 | 0.006632 | 52.821 | 3.65e-13 |
| hip OA | rs79056043 | 0.07920 | 0.010910 | 52.681 | 3.92e-13 |
| hip OA | rs6556077 | -0.04090 | 0.005640 | 52.581 | 4.13e-13 |
| hip OA | rs11021409 | -0.05050 | 0.007032 | 51.573 | 6.90e-13 |
| hip OA | rs849139 | -0.04030 | 0.005628 | 51.282 | 8.00e-13 |
| hip OA | rs35997606 | -0.04250 | 0.005935 | 51.281 | 8.00e-13 |
| hip OA | rs10974438 | -0.03930 | 0.005519 | 50.715 | 1.07e-12 |
| hip OA | rs509236 | 0.03990 | 0.005625 | 50.308 | 1.31e-12 |
| hip OA | rs833061 | -0.03840 | 0.005569 | 47.543 | 5.38e-12 |
| hip OA | rs876531 | -0.03700 | 0.005377 | 47.349 | 5.94e-12 |
| hip OA | rs9436635 | -0.05330 | 0.007790 | 46.820 | 7.78e-12 |
| hip OA | rs113799112 | 0.09000 | 0.013180 | 46.653 | 8.47e-12 |
| hip OA | rs17566944 | -0.04250 | 0.006235 | 46.455 | 9.37e-12 |
| hip OA | rs6501201 | 0.04110 | 0.006054 | 46.089 | 1.13e-11 |
| hip OA | rs11001788 | 0.03730 | 0.005509 | 45.848 | 1.28e-11 |
| hip OA | rs2616559 | -0.08190 | 0.012170 | 45.278 | 1.71e-11 |
| hip OA | rs459094 | 0.03910 | 0.005823 | 45.080 | 1.89e-11 |
| hip OA | rs67641990 | -0.04060 | 0.006080 | 44.588 | 2.43e-11 |
| hip OA | rs12040949 | -0.03620 | 0.005460 | 43.953 | 3.36e-11 |
| hip OA | rs75686861 | 0.05980 | 0.009036 | 43.797 | 3.64e-11 |
| hip OA | rs4073717 | -0.04330 | 0.006572 | 43.415 | 4.43e-11 |
| hip OA | rs3861871 | 0.03490 | 0.005312 | 43.168 | 5.02e-11 |
| hip OA | rs2836949 | 0.03450 | 0.005274 | 42.788 | 6.10e-11 |
| hip OA | rs1228024 | -0.03820 | 0.005863 | 42.447 | 7.26e-11 |
| hip OA | rs62233158 | -0.05770 | 0.008900 | 42.027 | 9.00e-11 |
| hip OA | rs3020418 | -0.03720 | 0.005742 | 41.979 | 9.22e-11 |
| hip OA | rs13165509 | 0.04120 | 0.006417 | 41.216 | 1.36e-10 |
| hip OA | rs3740129 | 0.03420 | 0.005342 | 40.981 | 1.54e-10 |
| hip OA | rs4836390 | 0.04690 | 0.007338 | 40.852 | 1.64e-10 |
| hip OA | rs11631921 | 0.03590 | 0.005622 | 40.781 | 1.70e-10 |
| hip OA | rs2119260 | -0.03840 | 0.006092 | 39.733 | 2.91e-10 |
| hip OA | rs8062183 | 0.03350 | 0.005323 | 39.602 | 3.11e-10 |
| hip OA | rs540864 | -0.04500 | 0.007162 | 39.477 | 3.32e-10 |
| hip OA | rs2731556 | -0.03400 | 0.005424 | 39.288 | 3.66e-10 |
| hip OA | rs725127 | -0.03440 | 0.005514 | 38.916 | 4.42e-10 |
| hip OA | rs3824488 | -0.05460 | 0.008764 | 38.816 | 4.66e-10 |
| hip OA | rs572543580 | 0.06760 | 0.010910 | 38.389 | 5.80e-10 |
| hip OA | rs11078685 | 0.03460 | 0.005592 | 38.290 | 6.10e-10 |
| hip OA | rs34811474 | -0.04040 | 0.006537 | 38.191 | 6.41e-10 |
| hip OA | rs9919834 | -0.08610 | 0.014010 | 37.794 | 7.86e-10 |
| hip OA | rs9986685 | -0.03450 | 0.005650 | 37.281 | 1.02e-09 |
| hip OA | rs12661964 | 0.06990 | 0.011450 | 37.265 | 1.03e-09 |
| hip OA | rs820780 | 0.03380 | 0.005544 | 37.171 | 1.08e-09 |
| hip OA | rs2147338 | -0.03220 | 0.005320 | 36.636 | 1.42e-09 |
| hip OA | rs73450750 | 0.04250 | 0.007044 | 36.406 | 1.60e-09 |
| hip OA | rs75495843 | -0.09450 | 0.015680 | 36.300 | 1.69e-09 |
| hip OA | rs79920061 | 0.06480 | 0.010770 | 36.217 | 1.76e-09 |
| hip OA | rs6020421 | -0.03340 | 0.005561 | 36.075 | 1.90e-09 |
| hip OA | rs3110823 | -0.04220 | 0.007031 | 36.027 | 1.95e-09 |
| hip OA | rs10790077 | 0.03240 | 0.005399 | 36.007 | 1.97e-09 |
| hip OA | rs7899657 | -0.03150 | 0.005259 | 35.877 | 2.10e-09 |
| hip OA | rs13030665 | 0.03190 | 0.005335 | 35.755 | 2.24e-09 |
| hip OA | rs76457470 | 0.07590 | 0.012810 | 35.115 | 3.11e-09 |
| hip OA | rs13264164 | -0.04580 | 0.007740 | 35.017 | 3.27e-09 |
| hip OA | rs2874496 | 0.03860 | 0.006526 | 34.986 | 3.32e-09 |
| hip OA | rs71396157 | 0.03910 | 0.006635 | 34.732 | 3.78e-09 |
| hip OA | rs12495734 | -0.03400 | 0.005778 | 34.620 | 4.01e-09 |
| hip OA | rs112635299 | -0.11490 | 0.019560 | 34.491 | 4.28e-09 |
| hip OA | rs3817428 | 0.03440 | 0.005866 | 34.385 | 4.52e-09 |
| hip OA | rs941123 | 0.03600 | 0.006145 | 34.318 | 4.68e-09 |
| hip OA | rs12197849 | -0.04160 | 0.007118 | 34.154 | 5.09e-09 |
| hip OA | rs35164552 | -0.04100 | 0.007031 | 34.003 | 5.50e-09 |
| hip OA | rs7294671 | 0.03060 | 0.005253 | 33.934 | 5.70e-09 |
| hip OA | rs12919887 | 0.03590 | 0.006197 | 33.559 | 6.92e-09 |
| hip OA | rs2071358 | 0.03830 | 0.006615 | 33.525 | 7.04e-09 |
| hip OA | rs2638281 | -0.03600 | 0.006245 | 33.232 | 8.18e-09 |
| hip OA | rs111400190 | -0.03920 | 0.006801 | 33.221 | 8.22e-09 |
| hip OA | rs1977810 | -0.04040 | 0.007026 | 33.061 | 8.93e-09 |
| hip OA | rs146011609 | -0.04860 | 0.008457 | 33.025 | 9.10e-09 |
| hip OA | rs4795574 | -0.03840 | 0.006684 | 33.004 | 9.20e-09 |
| hip OA | rs883921 | 0.03210 | 0.005589 | 32.983 | 9.30e-09 |
| hip OA | rs10264916 | 0.03360 | 0.005854 | 32.949 | 9.46e-09 |
| hip OA | rs17765868 | -0.03070 | 0.005353 | 32.895 | 9.73e-09 |
| hip OA | rs2946379 | -0.03280 | 0.005732 | 32.739 | 1.05e-08 |
| hip OA | rs942233 | 0.03260 | 0.005740 | 32.261 | 1.35e-08 |
| hip OA | rs116346582 | -0.04620 | 0.008161 | 32.044 | 1.51e-08 |
| hip OA | rs13084180 | 0.04240 | 0.007498 | 31.978 | 1.56e-08 |
| hip OA | rs2566752 | 0.03070 | 0.005435 | 31.905 | 1.62e-08 |
| hip OA | rs2378792 | 0.03500 | 0.006205 | 31.818 | 1.69e-08 |
| hip OA | rs2669046 | -0.02990 | 0.005303 | 31.791 | 1.72e-08 |
| hip OA | rs4734233 | -0.03150 | 0.005611 | 31.516 | 1.98e-08 |
| hip OA | rs1997958 | 0.03410 | 0.006096 | 31.292 | 2.22e-08 |
| hip OA | rs2836455 | 0.03830 | 0.006852 | 31.245 | 2.27e-08 |
| hip OA | rs4566248 | -0.03420 | 0.006178 | 30.645 | 3.10e-08 |
| hip OA | rs1749850 | -0.03140 | 0.005703 | 30.319 | 3.67e-08 |
| hip OA | rs7170658 | -0.03250 | 0.005917 | 30.166 | 3.97e-08 |
| hip OA | rs12534570 | -0.03220 | 0.005886 | 29.925 | 4.49e-08 |

**Supplementary Table S3.** Summary F-statistics across SNPs per exposure–outcome pair.

| Exposure | Outcome | # SNPs | Mean F | Median F | Min F | Max F |
| --- | --- | --- | --- | --- | --- | --- |
| neck | knee | 7 | 35.76180 | 31.41840 | 29.94272 | 47.03304 |
| neck | hand | 7 | 35.76180 | 31.41840 | 29.94272 | 47.03304 |
| neck | hip | 7 | 35.76180 | 31.41840 | 29.94272 | 47.03304 |
| knee | neck | 128 | 46.03315 | 38.67571 | 29.91031 | 164.18113 |
| hand | neck | 18 | 42.00912 | 37.11122 | 30.21203 | 94.89358 |
| hip | neck | 128 | 54.83486 | 42.61731 | 29.92450 | 199.48376 |

**Supplementary Figure S1.**
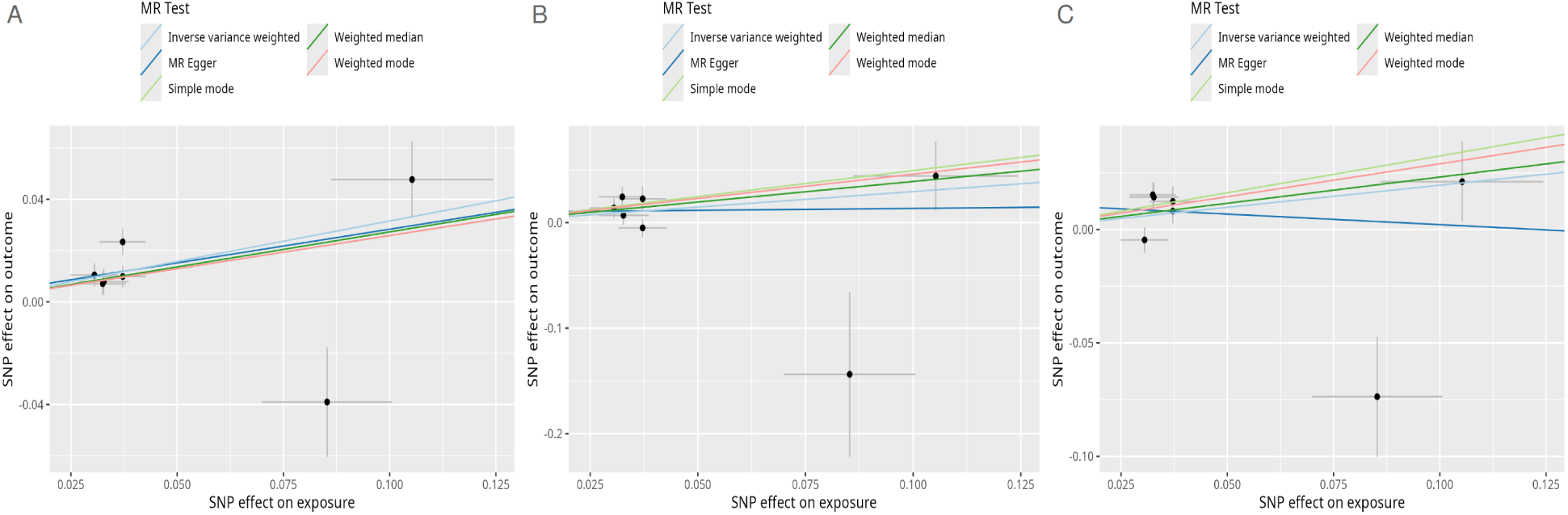
Scatter plots showing the effect of genetically predicted neck pain on knee osteoarthritis (A), hand osteoarthritis (B), and hip osteoarthritis (C)

**Supplementary Figure S2.**
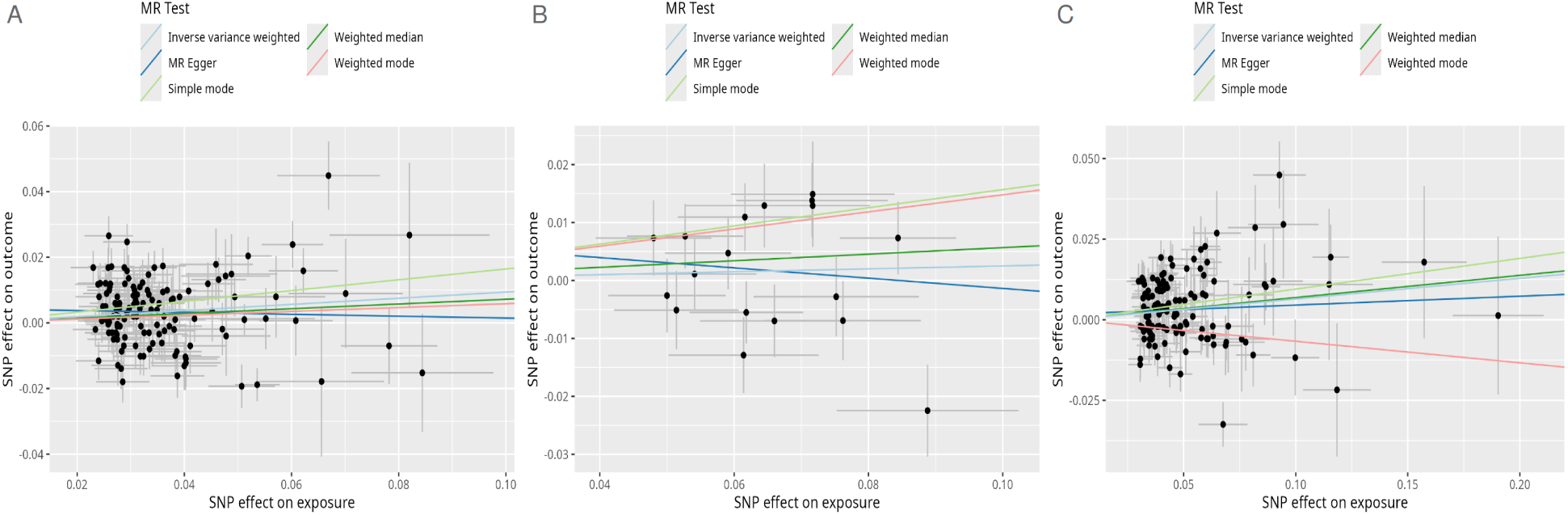
Scatter plots showing the effect of genetically predicted knee osteoarthritis (A), hand osteoarthritis (B), and hip osteoarthritis (C) on neck pain.

**Supplementary Figure S3.**
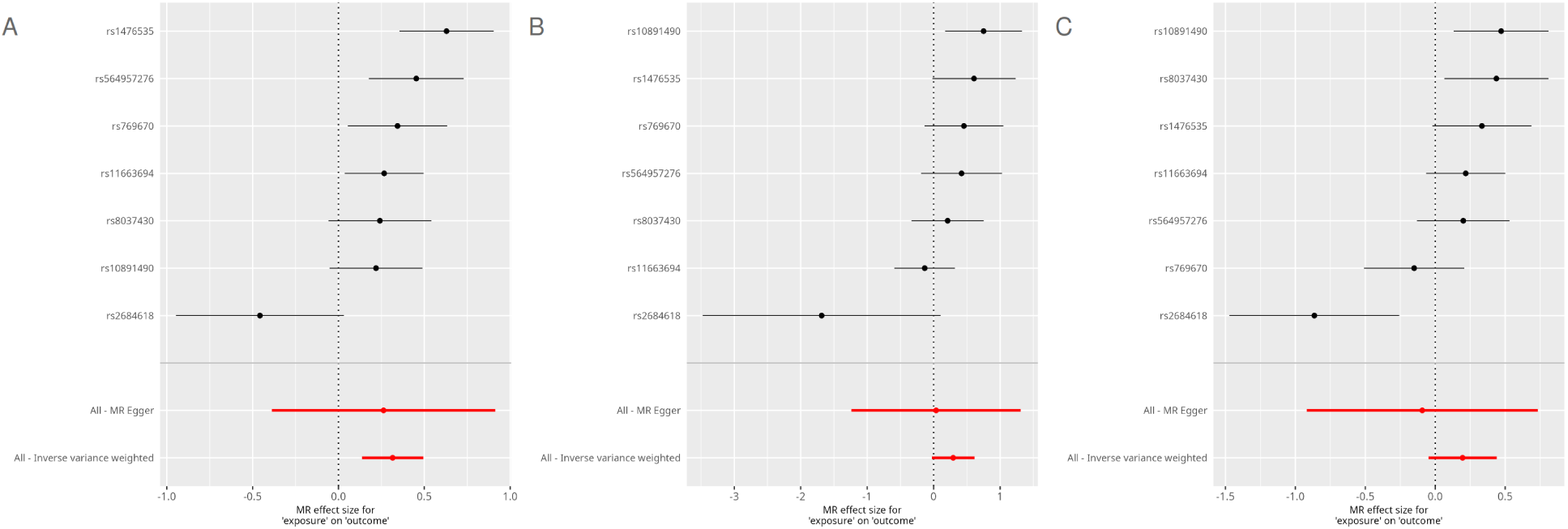
Per-SNP forest showing the effect of genetically predicted neck pain on knee osteoarthritis (A), hand osteoarthritis (B), and hip osteoarthritis (C).

**Supplementary Figure S4.**
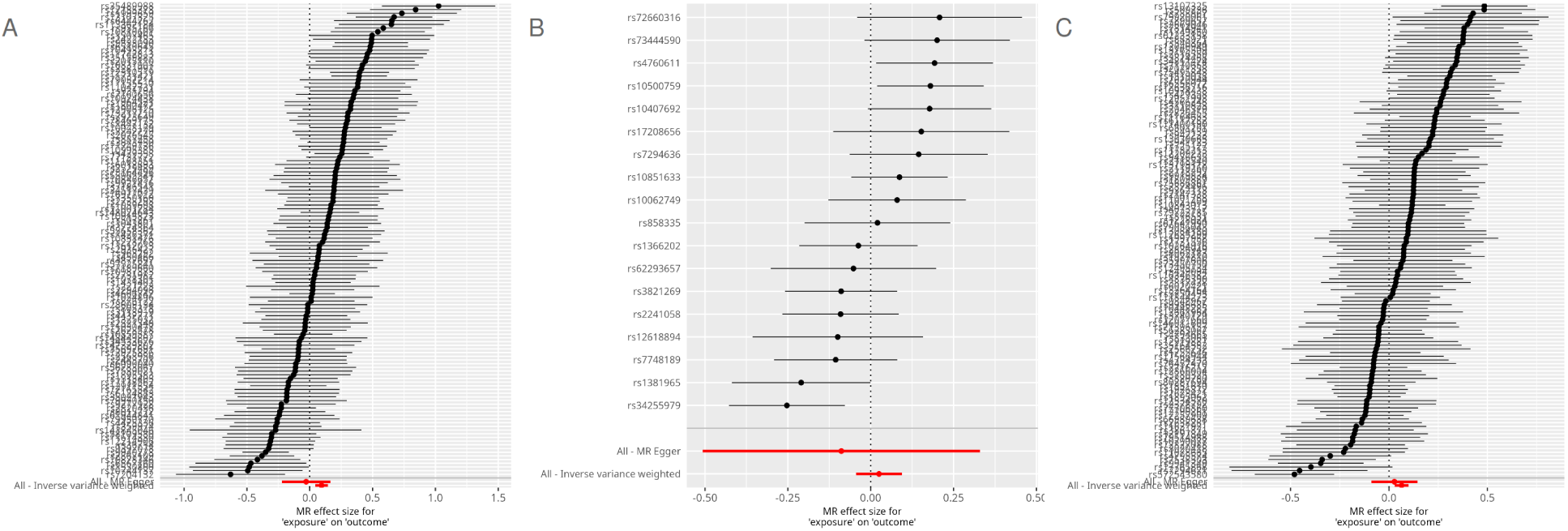
Per-SNP forest plots from Mendelian randomization assessing the effect of genetically predicted knee osteoarthritis (A), hand osteoarthritis (B), and hip osteoarthritis (C) on neck pain.

**Supplementary Figure S5.**
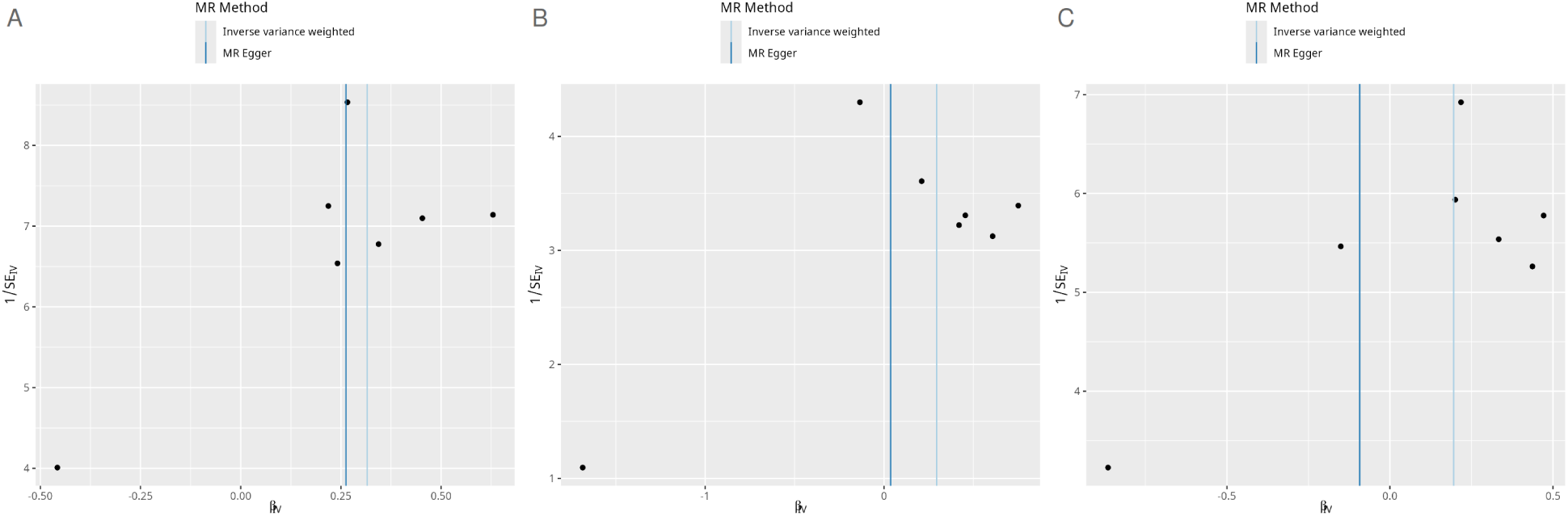
Funnel plots from Mendelian randomization assessing the effect of genetically predicted neck pain on knee osteoarthritis (A), hand osteoarthritis (B), and hip osteoarthritis (C).

**Supplementary Figure S6.**
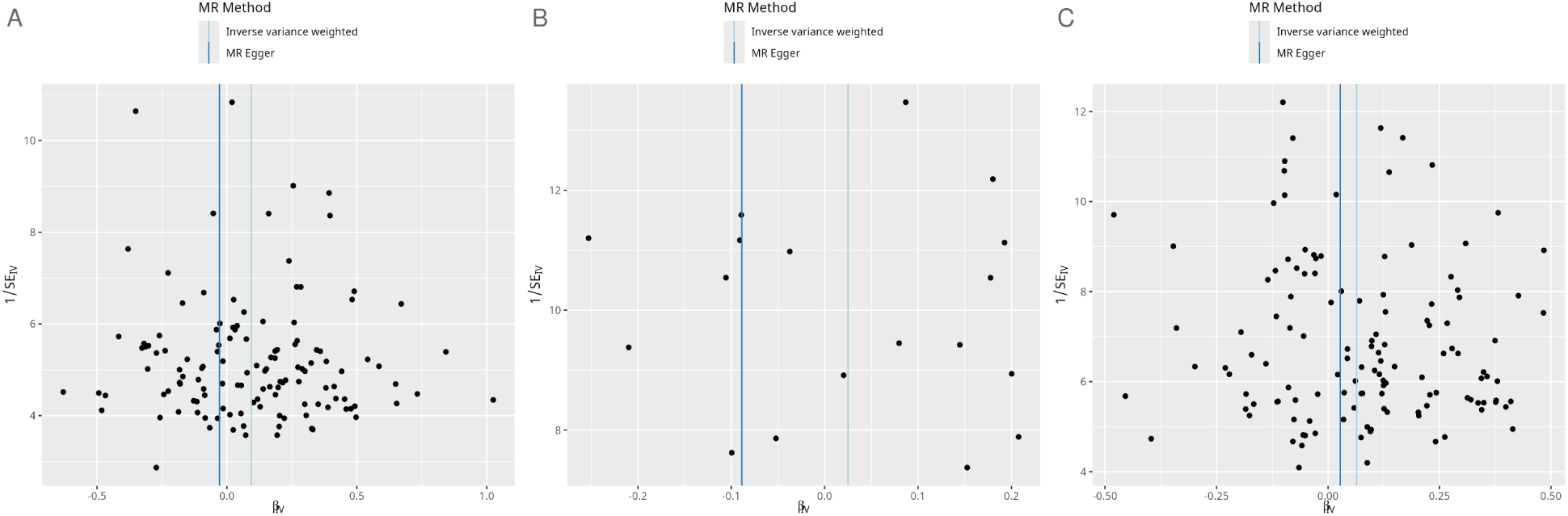
Funnel plots from Mendelian randomization assessing the effect of genetically predicted knee osteoarthritis (A), hand osteoarthritis (B), and hip osteoarthritis (C) on neck pain.

**Supplementary Figure S7.**
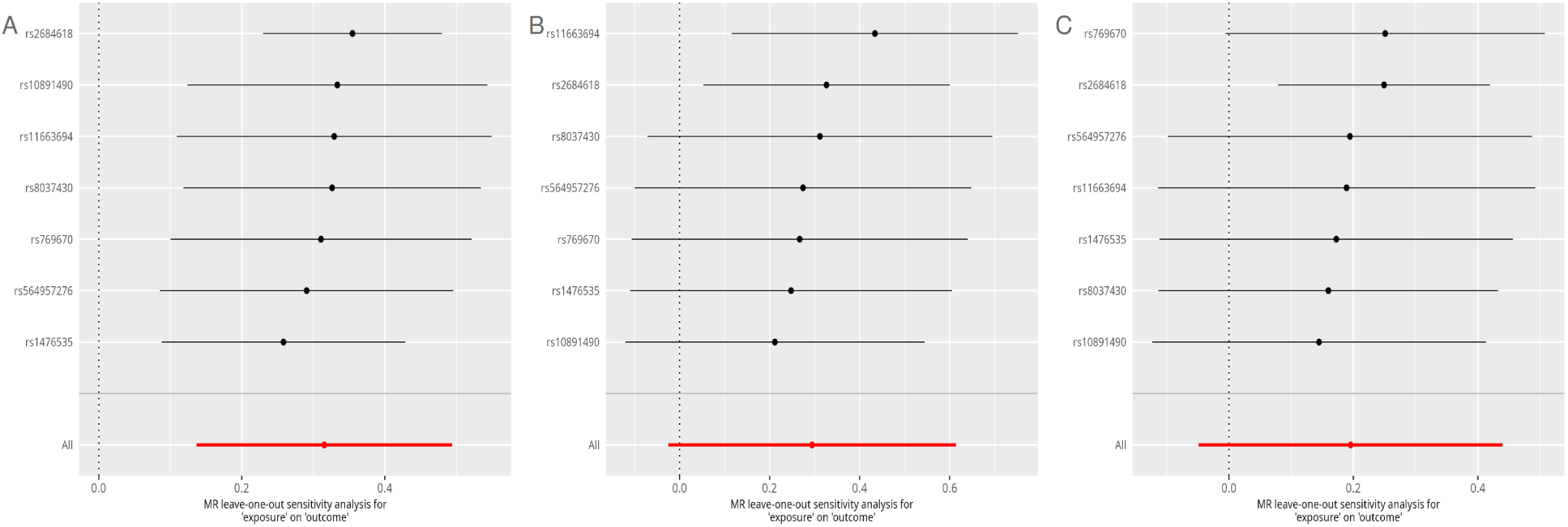
Leave-one-out analyses from Mendelian randomization assessing the effect of genetically predicted neck pain on knee osteoarthritis (A), hand osteoarthritis (B), and hip osteoarthritis (C).

**Supplementary Figure S8.**
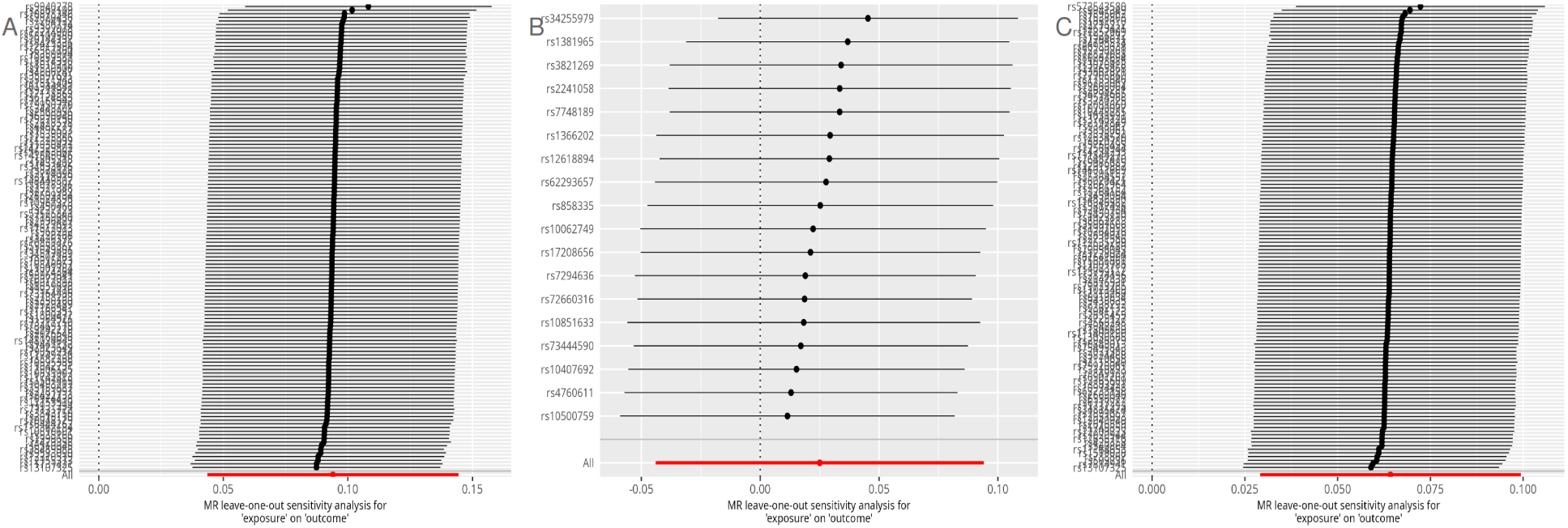
Leave-one-out analyses from Mendelian randomization assessing the effect of genetically predicted osteoarthritis on neck pain: (A) knee osteoarthritis, (B) hand osteoarthritis, (C) hip osteoarthritis.

